# Impact of increased diagnosis for early HIV infection and immediate antiretroviral treatment initiation on HIV transmission among men who have sex with men in the Netherlands

**DOI:** 10.1101/2024.04.10.24305619

**Authors:** Alexandra Teslya, Janneke Cornelia Maria Heijne, Maarten Franciscus Schim van der Loeff, Ard van Sighem, Jacob Aiden Roberts, Maartje Dijkstra, Godelieve J. de Bree, Axel Jeremias Schmidt, Kai J. Jonas, Mirjam E. Kretzschmar, Ganna Rozhnova

**Affiliations:** Julius Center for Health Sciences and Primary Care, University Medical Center Utrecht, Utrecht University, Utrecht, The Netherlands; Department of Infectious Diseases, Public Health Service of Amsterdam, Amsterdam, The Netherlands; Department of Internal Medicine, Division of Infectious Diseases, Amsterdam Institute for Immunology and Infectious Diseases, Amsterdam UMC, University of Amsterdam, Amsterdam, The Netherlands; Amsterdam UMC location University of Amsterdam, Amsterdam Public Health Research Institute, Amsterdam, The Netherlands; Stichting HIV Monitoring, Amsterdam, The Netherlands; Sigma Research, Department of Public Health, Environments and Society, London School of Hygiene and Tropical Medicine, London, United Kingdom; Medicine and Health Policy Unit, German AIDS Federation, Berlin, Germany; Faculty of Psychology and Neuroscience, Maastricht University, Maastricht, The Netherlands; Center for Complex Systems Studies (CCSS), Utrecht University, Utrecht, The Netherlands; BioISI – Biosystems & Integrative Sciences Institute, Faculdade de Ciências, Universidade de Lisboa, Lisbon, Portugal; Faculdade de Ciências, Universidade de Lisboa, Lisbon, Portugal

## Abstract

The number of new HIV infections among men who have sex with men (MSM) in the Netherlands has been decreasing, but additional efforts are required to bring it further down. This study aims to assess the impact of increased diagnosis for early HIV infection combined with immediate antiretroviral treatment (ART) initiation on reducing HIV transmission among MSM. We developed an agent-based model calibrated to HIV surveillance and sexual behavior data for MSM in the Netherlands. We simulated a 10-year intervention that accelerates HIV diagnosis during the first 3 or 6 months after HIV acquisition across five levels of increased diagnosis rates (2, 4, 8, 16, and 32-fold), followed by immediate ART initiation. The upper limit of the intervention’s impact over 10 years is projected to lower median cumulative HIV infections from 469 (interquartile range [IQR]: 300–681), projected without the intervention, to 184 (IQR: 142-–239), denoted as maximum impact. A 16-fold increase in the diagnosis rate within 3 months after HIV acquisition results in 263 (IQR: 182–349) infections. Further increases in the diagnosis rate show diminishing returns, failing to reach the maximum impact. By extending the scope of the intervention to individuals who acquired HIV infection within the previous 6 months, a 16-fold increase in the diagnosis rate approaches closely the maximum impact of the intervention. Accelerating early HIV diagnosis through increased awareness, screening, and testing can further reduce transmission among MSM, provided diagnosis rates rise significantly. Meeting this goal necessitates a stakeholder needs assessment.

**Author summary:** In recent years, in the Netherlands, the annual number of new HIV infections in the population of men who have sex with men (MSM) has been declining. Using an agent-based model calibrated to the current state of the HIV epidemic in MSM in the Netherlands, we explored the potential impact of an intervention that accelerates diagnosis in individuals with early HIV infection and facilitates immediate antiretroviral treatment initiation on further reducing the number of new HIV infections. Our projections indicate that such an intervention can noticeably reduce onward HIV transmission in the population of MSM. To achieve this, the diagnosis rate for individuals with early HIV infection would need to increase 16-fold. Increasing the diagnosis rate beyond this level is expected to bring marginal improvements, without approaching the maximum potential of the intervention. Extending the intervention to target individuals who acquired HIV within the previous 6 months could further lower the number of new HIV infections, bringing it closer to the maximum impact. Therefore, the intervention, which achieves an increased diagnosis rate in individuals with early HIV who then immediately initiate ART, can bring forth substantial reductions in new HIV infections, even against a backdrop of an already declining trend of the HIV epidemic.

## Introduction

In recent years, the Netherlands has made significant progress in containing the HIV epidemic among men who have sex with men (MSM). This success is evident in observable epidemic trends, such as the decreasing numbers of annual new HIV diagnoses [1] and can be attributed to effective care and prevention programs aligning closely with the 95-95-95 UNAIDS goals set for 2025 [2]. The goals define the percentages of diagnosed individuals out of the total population with HIV, individuals using antiretroviral treatment (ART) out of the total diagnosed population, and virally suppressed individuals out of the total population on ART, collectively referred to as the cascade of care. While the UNAIDS 95-95-95 goals currently stand at 96-97-97 for the overall MSM population in the Netherlands [1], HIV transmission is still ongoing within this group [3].

In 2022, MSM remained one of the key populations, accounting for more than 54% of new HIV diagnoses in the Netherlands [1]. The estimated annual number of newly acquired HIV infections among MSM has been steadily decreasing from 660 (CI: 620–720) in 2010 to 80 (CI: 40–150) in 2022 [1]. To reach the goal of zero new HIV infections, intervention efforts must be scaled up, and additional measures might be required to further reduce onward transmission. On one hand, addressing late diagnosis and care initiation remains an important goal. To wit, in 2022, a substantial proportion of new HIV diagnoses in MSM occurred in late and advanced stages of HIV infection, with 34% and 21% of all diagnoses, respectively [1]. On the other hand, a large proportion of HIV transmissions originated from individuals who had acquired HIV infection within the preceding 12 months [4]. Thus, increasing the diagnosis rate of MSM with early HIV infection and swiftly linking them to care to achieve an undetectable viral load is crucial. This strategy effectively will shorten the period of infection transmission and is paramount for achieving HIV elimination. Importantly, it will also prevent the decline of CD4 cell counts and subsequently halt the progression of health deterioration in MSM with HIV.

In the early 2000s, ART initiation was guided by the declining count of CD4 cells in people with HIV. In 2015, in response to the INSIGHT trial results [5], a new recommendation advocating for early treatment initiation, regardless of CD4 count, was adopted. The study by Kroon et al. [6] estimated that immediate ART initiation by individuals who were diagnosed in the acute stage of HIV infection could reduce the projected number of new HIV infections by approximately a factor of 5, compared to the scenario without early detection and immediate ART initiation. A systematic review and meta-analysis by Ford et al. [7] concluded that ART initiation on the day of diagnosis increases the odds of achieving suppression and remaining in care after 1 year. Zhao et al. [8] observed that a decrease in the time between HIV diagnosis and ART initiation was associated with increased odds of achieving viral suppression within 1 year of initiation. Thus, decreasing the time between diagnosis and ART initiation could contribute to a reduction in onward transmission.

Targeted screening for early HIV infection and immediate ART initiation among MSM was demonstrated to be a feasible and effective strategy at the Amsterdam sexually transmitted infection clinic [9]. The strategy involved a media campaign aiming to increase awareness of early HIV infection among MSM [10], a rapid diagnostic test for early infection, and a same-day referral trajectory to initiate ART. In this strategy, MSM with possible early infection, identified using a validated early-infection risk score, received point-of-care HIV-RNA testing. MSM diagnosed with early infection were immediately referred to start ART within 24 hours. This approach resulted in increased early HIV infection diagnoses and reduced the time from diagnosis to viral suppression [9]. A national rollout of such a strategy could move us closer to eliminating the HIV epidemic among MSM in the Netherlands. However, for the effective implementation of this intervention at the population level, it is essential to determine the optimal parameters, including the specific increase needed in the diagnosis rate and the precise target populations. While investigating the population-level effects of this strategy empirically is challenging, mathematical models can be used to determine its potential as a public health intervention.

The importance of early HIV infection in efforts to eliminate HIV transmission has been a subject under consideration for well over a decade [11]. Recent modeling studies have demonstrated that accelerated diagnosis of early HIV infection can be a successful strategy in reducing transmission among MSM. A study set in Peru, a region with a notably high rate of new HIV infections, projected that a significant number of new HIV infections can be prevented over 20 years by increasing the diagnosis rate and initiating ART early in individuals with early HIV infection and initiating ART [12]. Similar findings, in the context of the general MSM population in the USA, were shown in another modeling study where the increase in early HIV diagnosis and ART uptake led to notable reductions in the number of new HIV infections and a subsequent decrease in HIV prevalence over a two-decade span [13]. Moreover, a study focusing on MSM in San Francisco highlighted the importance of quicker ART initiation, showing that reducing the time from diagnosis to treatment initiation could significantly decrease the number of new HIV infections [14]. However, the effectiveness of accelerated initiation of ART in stemming onward HIV transmission may depend on the epidemic context, as evidenced by a modeling study by Kusejko et al. [15] in Switzerland which found that early initiation of ART resulted in a modest decrease in expected number of new HIV infections as compared to increasing HIV testing rates in individuals with early HIV infection [15]. Therefore, the combination of the two could be key to reducing onward HIV transmission.

Currently, there are no modeling studies evaluating the impact of an intervention aiming to accelerate the diagnosis of MSM with early HIV infection combined with immediate ART in the Netherlands. Building on the findings of the studies outlined above, we investigate the effectiveness of such an intervention in the Dutch setting. Our study aims to provide novel insights that could enhance the development of interventions aimed at advancing the elimination of HIV, particularly in settings with low rates of new HIV infections.

## Materials and methods

### Model overview

To project HIV transmission dynamics among MSM in the Netherlands, the developed agent-based model captures three types of processes: demographic dynamics, sexual network dynamics, and HIV disease progression and transmission dynamics. The demographic dynamics include the entrance of new individuals into the population, aging, and the exit of individuals from the population due to background mortality or cessation of sexual activity. The sexual network dynamics capture steady (long-term, non-overlapping with other steady partnerships) and non-steady (short-term, with the potential for concurrency) sexual partnerships that form and dissolve dynamically. HIV disease progression and transmission capture the history of natural HIV infection, and current practices of HIV care and prevention in the Netherlands, such as the cascade of care and pre-exposure prophylaxis (PrEP). The simulation advances one day at a time.

### Demography

The modeled population consisted of 25,000 individuals, scaled down from an estimated MSM population in the Netherlands of 200,000–300,000 individuals [16]. We considered sexually active individuals between 15 and 75 years of age. Individuals enter the population at the age of 15 years, age as the simulation advances, and exit the population once they reach 75 years. They can also leave the population as a result of background age-dependent mortality.

### Sexual network dynamics

In the context of MSM, anal intercourse is the predominant mode of transmission [17]. We model HIV transmission through anal intercourse in steady and non-steady sexual partnerships. Individuals can enter new partnerships, and existing partnerships can dissolve. A steady partnership is a long-term partnership. Individuals can have at most one steady partner at a time. All individuals in the population have the same propensity to enter a steady partnership. The term non-steady partnership designates all other partnerships where anal intercourse can take place. Non-steady partnerships are short-term and may be concurrent with other partnerships, both steady and non-steady. The propensity to enter new non-steady partnerships is an intrinsic property of an individual and depends on age and on being in a steady partnership. This property may change as individuals age, or when their steady-partner state changes. The formation of both types of partnerships is influenced by the effects of age and HIV status on selecting a partner, referred to as age assortativity and serosorting, respectively.

### HIV dynamics

We use a standard HIV framework for modeling the natural history of HIV infection, which is viewed as the sequence of early, chronic, and AIDS infection stages. In our study, we adopt an approach similar to that by Dijkstra et al. [9], where early infection lasts, on average, 89 days. This duration aligns with the cumulative duration of the first five stages of infection in the Fiebig classification [18] that delineates substages of early infection corresponding to distinct levels of HIV viremia, antibody seroconversion responses, and sensitivity of diagnostic methods needed to detect HIV infection. After an infection has occurred, individuals who acquired HIV progress through the early infection stage. Subsequently, individuals enter the chronic infection stage, which is an extended period where the potential to transmit HIV remains approximately constant and significantly lower than in the early infection stage. Once the chronic infection stage has concluded, individuals enter the AIDS stage, which we further divide into early and late AIDS stages. The early AIDS stage is characterized by an increased probability of transmitting HIV via anal intercourse compared to the chronic stage and additional mortality due to AIDS. Finally, a rare occurrence in the Netherlands, individuals who survive the early AIDS stage enter the late AIDS stage. This stage is characterized by a rapid rise in viral load, cessation of sexual activity, and increased mortality relative to the early AIDS stage.

### Baseline care and prevention

The model captures the current routine HIV care and prevention programs in the Netherlands, including diagnosis-treatment-suppression cascade and PrEP. Since September 2019, PrEP can be obtained through the national PrEP program and via a prescription from general practitioners. We model continuous PrEP use, resulting in a substantial reduction in the likelihood of HIV acquisition during anal intercourse with individuals with HIV. We approximate the eligibility criteria for PrEP by considering the number of sexual partners in the past 6 months. The rates of enrollment in and exit from the PrEP program are calibrated so that, following the initial years of the PrEP rollout, 5% of the population is using PrEP. Individuals who acquire HIV may be diagnosed, subsequently initiate ART, and eventually achieve viral suppression, rendering them effectively incapable of transmitting HIV. A fraction of individuals may discontinue ART or experience ART failure, returning to a state with an unsuppressed viral load. For simplicity, we assume that these individuals can be offered ART again.

### HIV transmission within partnerships

HIV transmission between two sexual partners via anal intercourse depends on the HIV infection stage of the partner with HIV, the effective sexual contact rate between the two partners, and the use of PrEP. The effective sexual contact rate can be further broken down into the frequency of anal intercourse acts and the probability of condom use during these acts. Condom use depends on the type of partnership and on PrEP use by individuals in the partnership and results from reconciling the personal, age-dependent preferences of individuals. We model condom use as constant for the entire duration of the partnership. The frequency of anal intercourse acts differs per type of partnership, with everyone modeled as having the same preference for the frequency of anal intercourse. Finally, based on existing evidence [19], we model that the effective sexual contact rates in serodiscordant partnerships where the partner with HIV is diagnosed are reduced compared to the baseline level of partnerships with unknown HIV status. We assumed the reduction to be by a factor of 2.

### Model parametrization and calibration

To accurately project HIV transmission dynamics in the MSM population in the Netherlands, we used available data to set model parameter values. Most parameter values were determined through direct calculation, known as parametrization, whereas others were derived by adjusting the model outputs to fit available data, a method called calibration. This approach was used for parameters that are particularly difficult to measure directly, such as the probability of transmission by an individual with chronic HIV infection during condomless anal intercourse. Additionally, although the diagnosis rates of individuals at various stages of HIV infection could be directly estimated from monitoring data, the model was calibrated with these estimates to accurately reflect the observed pattern of diagnosis rates in the MSM (men who have sex with men) population in the Netherlands. For estimating parameters relevant to sexual network dynamics and sexual behavior within partnerships, we utilized data collected from several sexual network/behavior surveys among MSM in the Netherlands, i.e., the cross-sectional European MSM Internet Survey 2017 (EMIS-2017) [20], the longitudinal Amsterdam Cohort Studies (ACS) [21], and the cross-sectional Amsterdam MSM Network study [22, 23]. To estimate HIV transmission dynamics, we used HIV surveillance data for MSM in the Netherlands [1, 24–30] and estimates from the literature [31–33]. Employing parametrization and calibration procedures, we obtained the main parameter set used throughout the analyses, for which the distance between the median of the model simulation and the HIV surveillance estimate/data [1] was minimized. For the selected parameter set, the simulation median always falls within the estimate of the confidence interval for the data. Sensitivity analyses were performed concerning the selected parameter values.

For a comprehensive description of parameter estimation, consult the S1 Appendix.

#### Parametrization

##### Demographic processes

We used data from Statistics Netherlands to calculate background age-dependent mortality rates and the age distribution of the MSM population. The latter was assumed to be similar to the age distribution of the general male population [34]. The entrance rate of new individuals is a Poisson-distributed process with its mean set such that the population size fluctuates around 25,000 individuals.

##### Sexual network

Using available data, we calculated the proportion of the population that has a steady partner and the switch rates of non-steady partners distributed by the age of individuals and the existence of a steady partner.

##### HIV transmission dynamics

Transition rates between HIV infection stages and the relative infectivity per contact in different HIV stages were parameterized using estimates from the literature [31–33].

#### Calibration

##### Sexual network

To obtain the rates of partnership formation and dissolution, we calibrated the model against the distribution of the rate of change of steady partners in the previous 12 months and the rate of change of non-steady partners in the previous 6 months using the data collected in EMIS-2017 [20] survey. To determine assortativity mixing and serosorting patterns, we calibrated the model against data collected in the Amsterdam MSM Network study [22, 23] and Amsterdam Cohort Studies [21].

##### HIV transmission dynamics

To estimate the baseline probability of HIV transmission per condomless anal intercourse, the diagnosis rates of individuals in different HIV stages, ART uptake, and viral suppression rates, we calibrated HIV transmission dynamics in the presence of currently realized standards of HIV care and prevention. The calibration targets were the estimated annual number of newly acquired HIV infections, the annual number of newly diagnosed HIV infections, the proportion of individuals diagnosed within specific time window after HIV acquisition (first 6 months, 6 to 12 months, and after more than 12 months), the proportion of individuals who have started treatment among those who received an HIV diagnosis, and the proportion of individuals who achieved a suppressed viral load among the total number of individuals who started treatment in the MSM population for the years 2017-2022 in the Netherlands [1, 26–30].

### Model scenarios

In all scenarios, at the beginning of the simulation, the population age structure is seeded using the state of the male population in 2014. We initiate HIV dynamics using the population distribution in the cascade of care in 2016 [25] and allow it to progress until 2023. Depending on the modeled scenario, an intervention may then be initiated and run for 10 years starting in 2023. We simulate the following scenarios:

1. **Baseline scenario.** No additional interventions are modeled, except for the PrEP program and the cascade of care using the standard of care in 2016, wherein ART initiation increasingly occurred soon after HIV diagnosis.
2. **Intervention scenario.** An intervention that facilitates an increased diagnosis for individuals with early HIV infection and immediate ART initiation would start in 2023. We considered scenarios with the following specifications:

a. **Maximum impact scenario.** In this scenario, we simulate that every individual, whether with newly acquired HIV infection or with HIV infection acquired within the previous 6 months, begins ART right at the point of HIV acquisition or at the start of the intervention, whichever comes first. We achieve this by setting the diagnosis rates high enough to ensure immediate ART initiation for all eligible individuals upon HIV acquisition or with the intervention’s initiation. Although this scenario may not be realistic, examining its outcomes helps us understand the maximum potential impact of the intervention which combines increased diagnosis in individuals with early HIV infection with immediate initiation of ART.
b. **Accelerated diagnosis in individuals with early HIV infection.** In this scenario, we explore incremental increases in the diagnosis rate for individuals with early HIV infection, combined with immediate initiation of ART. The diagnosis rates are increased relative to the baseline level by factors of 2, 4, 8, 16, and 32. At the baseline, 17% of individuals with newly acquired HIV are diagnosed during the early infection stage. As the diagnosis rates increase incrementally, the proportions of diagnosed individuals rise to 31%, 52%, 77%, 95%, and 100%, respectively.
c. **Accelerated diagnosis in individuals who acquired HIV within the previous 6 months.** This scenario captures the effect of an intervention that causes an increase in diagnosis rates not only in individuals with early HIV infection as in (b) but also in individuals who have entered the chronic HIV infection but have acquired HIV within the previous 6 months. Similarly, the diagnosis rate is increased (i.e., 2, 4, 8, 16, and 32-fold) compared to the baseline diagnosis rate, and individuals initiate ART on the same day.

### Model outcomes

To evaluate the intervention impact, we considered the annual and cumulative numbers of new HIV infections and new HIV diagnoses, the number of new HIV infections averted, as well as the distribution of the annual number of new HIV diagnoses by the duration between HIV acquisition and diagnosis. The cumulative numbers and the number of new HIV infections averted were calculated over 10 years starting from the intervention implementation on 1 January 2023. For each scenario, we generated 400 trajectories, summarizing the distribution of the model outputs using median values and inter-quartile ranges (IQR).

### Sensitivity analyses

We conducted sensitivity analyses for the model calibration with respect to the baseline probability of HIV transmission per contact, the rates of diagnosis of individuals with early infection, and individuals with chronic infection who acquired HIV in the previous 6 and 12 months, the rate of ART failure, as well as rates of ART initiation and viral load suppression. We also investigated the contribution of immediate initiation of ART on the impact of the intervention on HIV transmission dynamics. The full description of the sensitivity analyses is provided in the Subsections Sensitivity to parameter perturbation and Immediate initiation of ART in Section Additional Analyses in S2 Appendix.

## Results

### Model fit

The model fit to key data statistics of interest is shown in Figure 1. There was strong agreement between the model outputs and the data, namely the annual numbers of new HIV infections (Figure 1**a**) and new HIV diagnoses (Figure 1**b**). The model estimates of the proportions of individuals who received an HIV diagnosis within the first 6 and between 6 and 12 months following HIV acquisition marginally exceeded the data estimates (Figure 1**c**).

**Fig 1.**
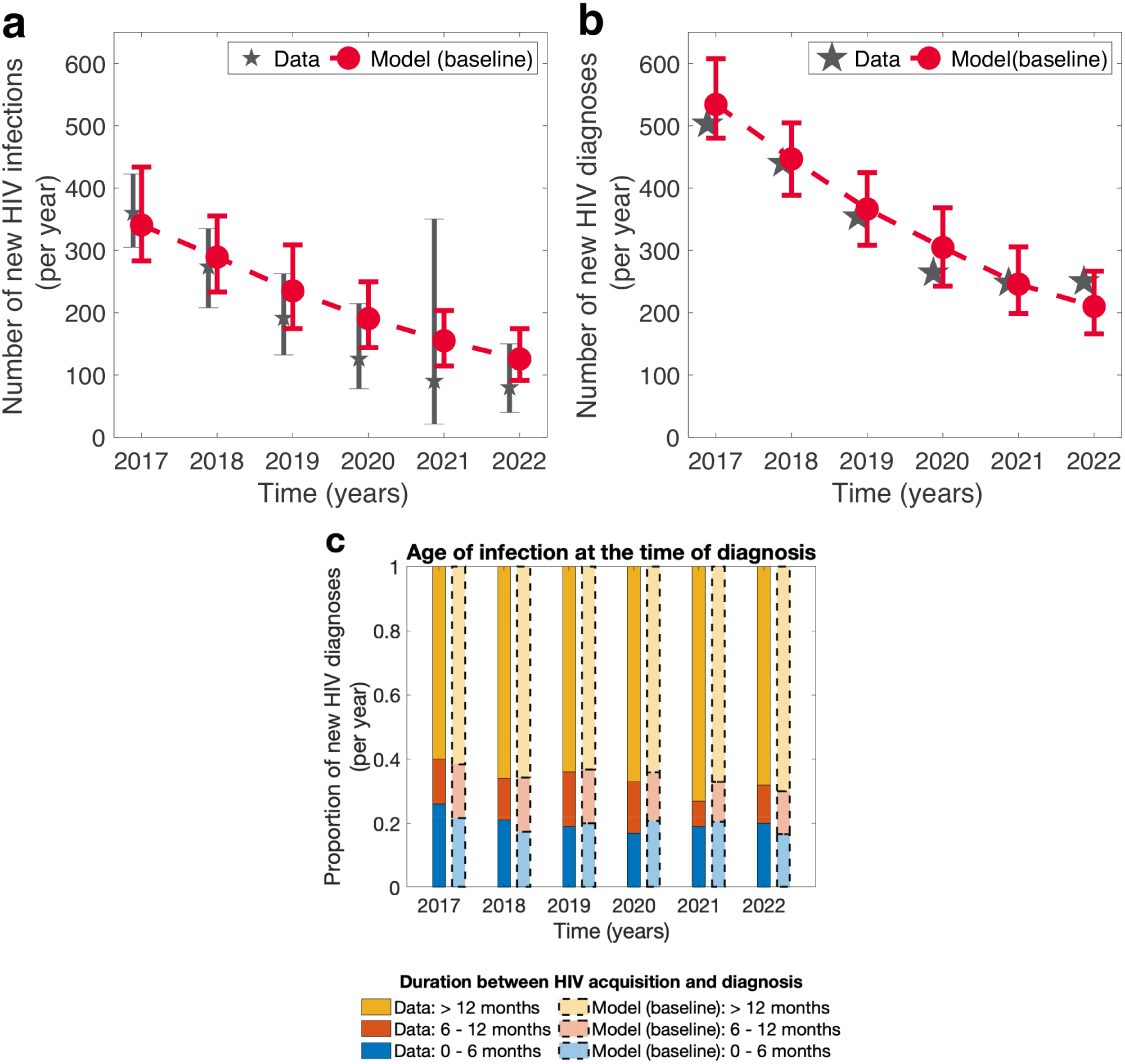
Model fit to the HIV surveillance data among MSM in the Netherlands. **a** Newly acquired HIV infections, **b** newly diagnosed HIV infections, and **c** distribution of the number of new HIV diagnoses by the duration between HIV acquisition and diagnosis for MSM, as estimated in the model and reported by the Stichting HIV Monitoring in the Netherlands. In **a** and **b**, the red dashed line depicts the baseline model output. For the data, the large grey stars and the bars denote the median estimates and the confidence intervals, respectively. For the model outputs, the large red dots and the bars denote the median estimates and the IQR, respectively. In **c**, the bar chart colors correspond to the time between HIV acquisition and diagnosis. For each year, bars with saturated colors and solid borders on the left correspond to the data, while the bars with less saturated colors and dashed borders on the right correspond to the model output.

### Maximum impact of the intervention

The comparison of the baseline model scenario with the maximum impact scenario is shown in Figures 2 and 3. Specifically, we examined the annual numbers of new HIV infections (Figure 2**a**) and new HIV diagnoses (Figure 2**b**). Furthermore, we assessed the distribution of the annual number of new HIV diagnoses by the duration between HIV acquisition and diagnosis to evaluate the intervention’s time frame of efficiency (Figure 3).

**Fig 2.**
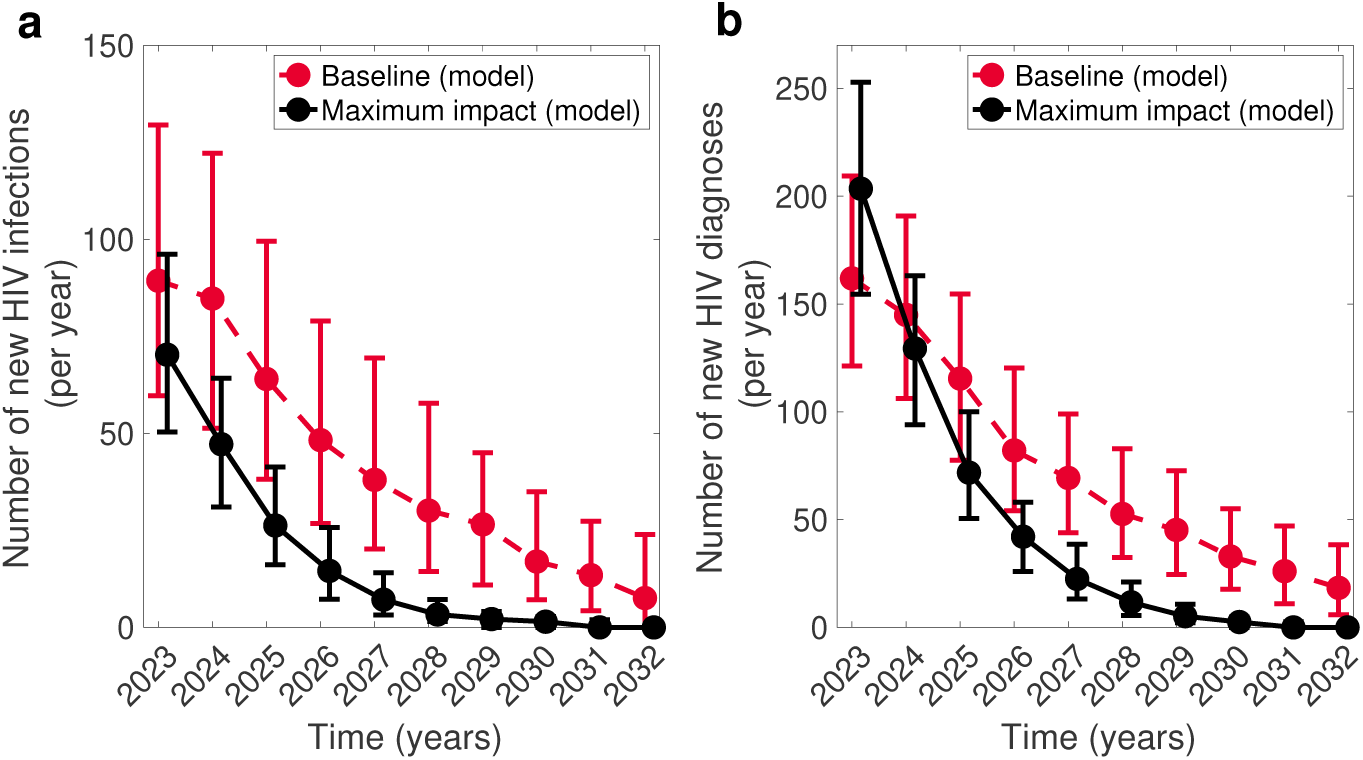
Model predictions for the maximum impact intervention. (**a**) Newly acquired HIV infections and (**b**) newly diagnosed HIV infections in the baseline and maximum impact model scenarios. The red dashed line depicts the baseline scenario without additional diagnosis of individuals with early infection. The black solid line depicts the maximum expected impact of an intervention where individuals with early infection are diagnosed at an increased rate compared to the baseline scenario and start ART immediately at the time point of receiving the diagnosis. The dynamics are summarized by the median estimates (circles) and the IQR (bars). The maximum impact intervention is expected to further accelerate the decline of the annual number of new infections. Importantly, the number of newly diagnosed individuals may drastically increase in the first year following the start of the intervention but is expected to decrease below the baseline level within two years.

**Fig 3.**
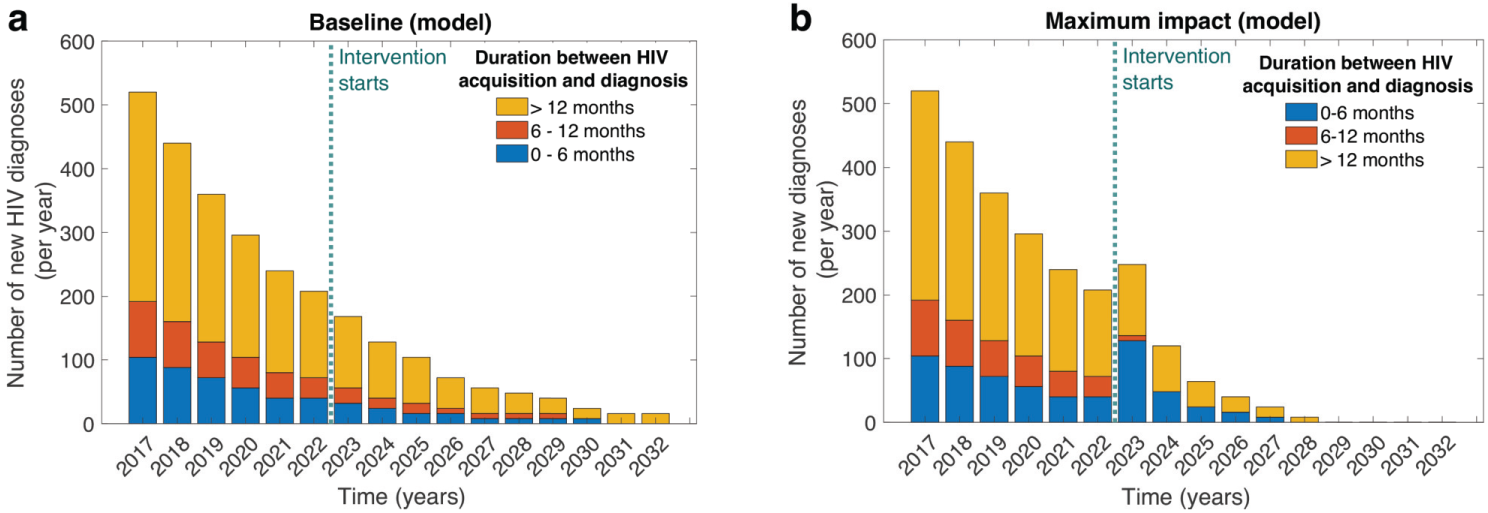
Distribution of new HIV diagnoses by the duration between HIV acquisition and diagnosis. (**a**) The baseline scenario without an increase in the diagnosis rate of individuals with early infection. (**b**) The maximum impact intervention combines an increased diagnosis rate of individuals with early infection with immediate ART initiation. Bar chart colors correspond to the duration between HIV acquisition and the time of diagnosis. The dotted teal line marks the start of the intervention. In the regime of maximum impact, six years after the start of the intervention, the number of diagnosed individuals was projected to be virtually zero.

#### New HIV infections

Our analyses indicated that in the context of continually decreasing low annual numbers of new HIV infections, as observed among MSM in the Netherlands, an intervention facilitating increased diagnosis of individuals with early HIV infection combined with immediate initiation of ART can further expedite this downward trend (Figure 2**a**). The effects of such intervention can become noticeable as early as the first year after implementation. In the baseline scenario, we anticipate the median annual number of new HIV infections to decline from 89 (IQR: 60–130) in 2023 to 8 (IQR: 0–24) in 2032. In the maximum impact scenario, we expect the median annual number of new HIV infections to decrease from 72 (IQR: 50–97) in 2023 to 0 (IQR: 0–1) in 2032, resulting in a cumulative decrease of 285 (IQR: 231–328) over 10 years.

#### New HIV diagnoses

The analysis of the annual number of new diagnoses (Figure 2**b**) indicated that, in the maximum impact scenario, there will be a sharp increase in the median number of newly diagnosed HIV infections from 161 (IQR: 90–189) at baseline to 203 (IQR: 155–253) in the year following the start of the intervention. However, the number of HIV diagnoses is expected to decrease to the baseline scenario level within the first two years of the intervention onset and subsequently drop significantly below it by the third year. This drastic decrease in newly diagnosed HIV infections signifies an overall reduction in HIV transmission induced by the intervention. In the maximum impact scenario, the model predicted a reduction in the median cumulative number of diagnoses from 753 (IQR: 558–1020) to 467 (IQR: 272–574) over 10 years.

#### Distribution of new HIV diagnoses by the duration between HIV acquisition and diagnosis

To analyze the window of efficiency of the maximum impact intervention, we examined the distribution of new HIV diagnoses by the duration between HIV acquisition and the time of diagnosis (Figure 3). In the baseline scenario, our model predicted that the majority of individuals continue to be diagnosed more than 12 months after HIV acquisition (Figure 3**a**). For the maximum impact intervention, the number and proportion of diagnoses in individuals with early HIV infection increased drastically in the first three years of the intervention (Figure 3**b**). Specifically, over the first three years, in the baseline scenario, a median of 72 (IQR: 40–120) out of 400 (IQR: 280–576) new diagnoses were in individuals who acquired HIV within the previous 6 months. In the maximum impact scenario, a median of 200 (IQR: 136–272) out of 432 (IQR: 304–584) individuals were expected to receive a diagnosis within the first 6 months following HIV acquisition. This number, owing to the decreased number of new HIV infections, reduced drastically in the subsequent years. In the maximum impact intervention scenario, no new HIV diagnoses were projected starting in 2029.

### Attaining maximum impact

To identify the increase that results in a reduction (relative to the baseline) of the number of new HIV infections comparable to that of the maximum impact intervention we investigated a range of diagnosis rates for individuals with early HIV infection (Figure 4). Additionally, we considered the impact of an extended intervention, where the diagnosis rate is increased in all individuals who acquired HIV infection within the previous 6 months. To perform these analyses, we calculated the cumulative number of new HIV infections (Figure 4**a**) and diagnoses (Figure 4**b**) over 10 years.

**Fig 4.**
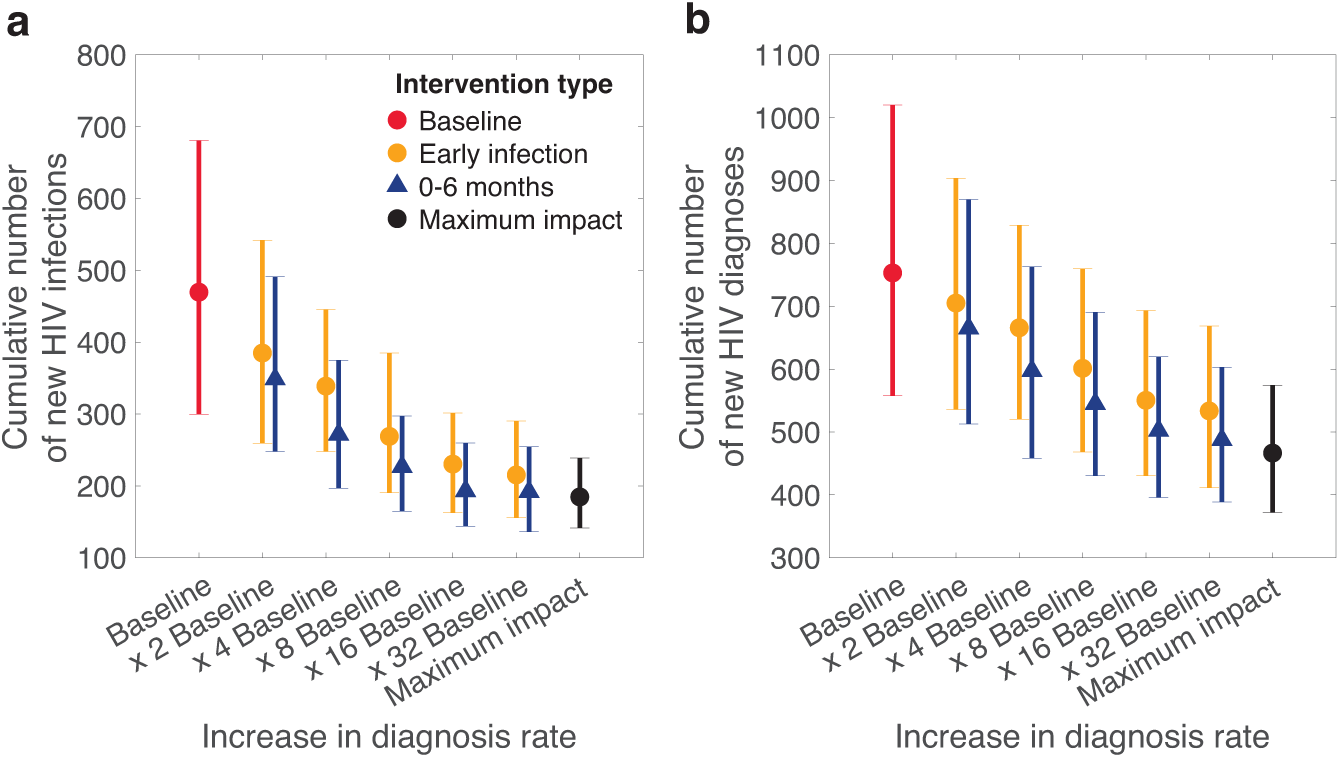
Impact of the intervention across different scenarios. (**a**) Cumulative HIV infections and (**b**) cumulative HIV diagnoses over 10 years. Red diamonds correspond to the baseline scenario without an additional increase in the diagnosis rates. Yellow dots and blue triangles correspond to the intervention where the diagnosis rate is increased, compared to the baseline, in individuals with early HIV infection or those who acquired HIV within the previous 6 months, respectively. The black dots mark the maximum impact scenario. In the regime of the HIV epidemic currently observed among MSM in the Netherlands, where the annual number of newly acquired HIV infections has been relatively low and decreasing, a 16-fold increase in the diagnosis rate during early HIV infection can lead to a substantial reduction in the appearance of new infections and, subsequently, new HIV diagnoses. However, modest increases in the diagnosis rate beyond this point do not succeed in achieving the maximum possible impact of the intervention and to reach it, a wider group of MSM needs to be engaged (i.e., those who acquired HIV within the previous 6 months). In such a case, the cumulative number of new HIV infections was projected to be lower than for the respective increase in diagnosing early infections only. Moreover, the maximum impact of the intervention can be achieved by a 16-fold increase in the diagnosis rate.

#### Accelerated diagnosis in individuals infected with early HIV infection

A 2-fold increase in the diagnosis rate of individuals with early HIV infection decreased the median cumulative infections from 469 (IQR: 300–681) at baseline to 385 (IQR: 260–542) and led to 84 total infections averted in 10 years, 30 of which occurred in the first five years (Figure 4**a**). An 8-fold increase in the diagnosis rate of early infections reduced the median cumulative infections to 236 (IQR: 191–385). The 8-fold increase had a more immediate impact than the 2-fold increase, with 200 infections averted in 10 years, 103 of which in the first five years and 97 in the following five years. A 16-fold increase reduced cumulative infections to 230 (IQR: 162–301), resulting in 239 total infections averted, 136 and 103 of which occurred in the first five years and the following five years, respectively. Finally, a 32-fold increase in the diagnosis rate did not yield a perceptible improvement relative to the 16-fold increase [i.e., 215 median cumulative infections (IQR: 156–290)]. Moreover, this outcome is not proximate to the maximum impact of the intervention [i.e., 255 infections averted (IQR: 152–421) and 191 infections averted]. Notably, in the first five years, the number of infections averted in the 32-increase intervention was almost equal to the 16-fold increase intervention [i.e., 141 (IQR: 74–199) vs 136 (IQR: 69–190) cumulative infections, respectively]. The trends we observed in the decrease in the cumulative number of new HIV infections were respectively reflected in the cumulative number of new HIV diagnoses (Figure 4**b**). Thus, an intervention that accelerates diagnosis in individuals with early HIV infection should aim for a sufficiently high increase in the diagnosis rate. However, once this threshold is reached, no additional benefit can be attained through further modest increases. Instead, consideration should be given to an additional expansion of the intervention involving a larger group of MSM with undiagnosed HIV infection.

#### Accelerated diagnosis in individuals infected within the previous 6 months

To identify such expansion, we investigated the outcomes of an intervention with an increased diagnosis rate in individuals who acquired HIV within the previous 6 months. We observed that this intervention yields a larger number of infections averted (Figure 4**a**, blue triangles) throughout the range of increases in the diagnosis rate that we considered. Moreover, the level of maximum impact can be achieved within the range of diagnosis rates that we investigated. For a 16-fold or larger increase in the diagnosis rate (Figure 4**a** and 4**b**, blue triangles), the maximum impact intervention predicted 184 median cumulative infections (IQR: 142–249), while the acceleration of diagnosis by the factor of 16 in individuals who acquired HIV within the previous 6 months predicted 193 infections (IQR: 144–260).

### Sensitivity analyses

We performed sensitivity analyses of the main findings with respect to a selected set of model parameters, specifically the baseline probability of HIV transmission per condomless anal intercourse, and the diagnosis rates of individuals in different HIV stages. We varied these parameters in a way that caused the annual number of new HIV infections and diagnoses in 2016–2022 to fluctuate about the calibration data and to follow a downward trend as seen in the HIV surveillance data in the Netherlands. We observed that while the projected cumulative number of new HIV infections and the cumulative number of new HIV diagnoses fluctuated around the outputs presented in the main analyses, the qualitative findings we reported in the main analysis held. More specifically, increased diagnosis in individuals with early HIV infection and in individuals who acquired HIV infection within the last 6 months caused a significant reduction in the number of new HIV infections over 10 years. An 16-fold increase in the diagnosis rate brought the impact of the intervention in terms of the cumulative numbers of new HIV infections and new HIV diagnoses in proximity to the maximal impact of the intervention.

Additionally, we explored the contribution of ART immediately after receiving an HIV diagnosis on the impact of the intervention. Repeating the analysis from the previous section, we examined the 10-year trends in both new HIV infections and diagnoses across various diagnosis rates among individuals with early HIV infection and those infected in the last 6 months. This time, individuals diagnosed through the intervention were assumed to start ART at the same rate as those identified via routine testing. Our findings suggest that, in our study context, under conditions similar to the contemporary Netherlands, when the increase in diagnosis rates is small, the added benefit of immediate ART initiation on reducing HIV incidence is not substantial beyond the impact of accelerated diagnosis. However, with larger increases in diagnosis rates, the immediate initiation of ART can result in averting a significant number of new HIV infections. However, even for large increases in the diagnosis rate, the contribution of ART is significantly smaller than that of the accelerated diagnosis. The detailed analyses are provided in the Subsections Sensitivity to parameter perturbation and Immediate initiation of ART in Section Additional Analyses in S2 Appendix.

## Discussion

In this study, we developed an agent-based model to investigate the potential impact of an intervention accelerating the diagnosis of MSM with early HIV infection, combined with immediate ART initiation, on the HIV epidemic among MSM in the Netherlands. Our analysis revealed that the intervention could significantly contribute to the decline in new HIV infections, particularly when diagnosis rates for early infection are thoroughly optimized. The model suggests that for individuals with early HIV infection, a 16-fold increase in the diagnosis rate, corresponding to 95% of new HIV infections diagnosed with early HIV, is optimal, with diminishing returns observed for modest decreases in terms of HIV infections averted beyond this threshold. Expanding the intervention to include individuals who have acquired HIV within the previous 6 months, an 8-fold increase in the diagnosis rate was shown to yield the same impact as when the intervention engaged only individuals with early HIV infection with a 16-fold increase in the diagnosis rate. Importantly, while our model captured increasing diagnosis rates combined with immediate ART for HIV infections acquired within the previous 6 months only, the real-world implementation of the intervention is likely to identify individuals with HIV infection older than 6 months as well. These individuals, upon being diagnosed through the intervention, would not be excluded from immediately receiving ART. This approach would result in a greater impact, as it captures and offers treatment to a broader spectrum of the HIV-infected population, not just those within the modeled 0-6 month period.

Our model aimed to accurately capture HIV transmission dynamics among MSM by portraying a realistic depiction of the dynamics of the sexual network among MSM. This portrayal was informed by data on partnership formation patterns and duration among MSM in the Netherlands. Key aspects relevant to HIV transmission, including serosorting, age assortativity, and condom use differentiated by partnership type and participant age, were integrated. We also modeled the standard of HIV care and prevention in the Netherlands. Furthermore, the model’s outputs were calibrated against relevant epidemiological statistics, specifically the incidence of new HIV diagnoses and the stage of infection at the time of HIV diagnosis, lending credibility to its projections and enhancing the trustworthiness of our findings.

Our model has several limitations, including the omission of the eclipse phase, the time between HIV acquisition and the first appearance of detectable viral RNA in plasma [35], and the simplification of care retention and prevention program progression. The impact of external factors, such as the COVID-19 pandemic, on sexual risk behavior also remains unaccounted for. Not accounting for these factors can affect the numerical accuracy of the models’ predictions by making the intervention appear more or less effective than it realistically can be expected under these conditions. For instance, not accounting for the eclipse stage when viral RNA is undetectable can lead to overestimation of the ability to diagnose individuals with early HIV infection. On the other hand, simplifying the details of care retention might lead to optimistic projections regarding the long-term engagement in the care of diagnosed individuals including those who were diagnosed via the early diagnosis intervention. These limitations highlight the importance of incorporating more detailed data on the eclipse phase and the nuances of care retention and prevention program progression for more accurate and actionable projections. While we acknowledge that not all MSM engage in anal intercourse by age 15 or at all, our model does not differentiate these behaviors, instead assuming that, under similar conditions, a uniform rate of anal intercourse among all individuals based on averaged data applies. This approach simplifies the modeling process, and while it may not capture the diverse sexual behaviors in the MSM population, it is designed to maintain an accurate average contact rate for the population studied. Together these factors might introduce some degree of uncertainty to the precise quantification of the intervention’s impact, but it is essential to note that our model accurately reproduces key epidemiological statistics central to our analysis, both numerically and in terms of dynamical trends. Consequently, we are confident that the core insights and directional trends predicted by our model will remain valid in similar HIV transmission contexts, as pertains to the low decreasing number of new HIV infections and a cascade of care proximate to UNAIDS’ 95-95-95 targets.

Inspired by targeted screening strategy for early HIV infection and immediate ART initiation among MSM at the Amsterdam sexually transmitted infection clinic [9], which focused on increasing early infection diagnosis through awareness campaigns combined with a robust HIV-risk score, same-day diagnosis, and immediate ART initiation, our study demonstrates the potential for expanding such strategies on a national scale. However, their success depends on the local epidemiological and social landscapes, underscoring the need for context-specific adaptations.

Mathematical modeling can elucidate how to adjust these interventions for varying contexts to optimize their impact, capturing diverse epidemiological profiles and sexual behaviors to tailor public health strategies. To effectively implement the interventions that our model explored, a comprehensive needs assessment will be essential. This assessment can identify barriers to engaging with interventions, such as access to information, the ability to undergo screening and HIV testing promptly, and the initiation and adherence to ART treatment. Factors including place of residence, age, and socio-demographic status significantly influence these aspects [36]. Our model primarily aimed at assessing the potential impact of increased diagnosis rates on reducing HIV transmission among MSM. As such, it directly focused on the increase in diagnosis rates without capturing the corresponding increase in screening and testing that such an intervention requires. Understanding the full scope of the intervention’s impact requires examining the intricacies of testing and screening processes, as well as considering the challenges in engaging the target population to achieve a necessary increase in diagnosis rates.

The success of targeted screening and immediate ART initiation in Amsterdam, supported by media campaigns developed with the MSM community, underscores the potential of such approaches [10]. Yet, the effectiveness of replicating these strategies in regional contexts different from Amsterdam highlights the complexity of adapting interventions to different cultural landscapes. For instance, the original campaign’s impact in Amsterdam facilitated through collaboration with community stakeholders and tailored media strategies, may not directly translate to regions with distinct cultural and socio-demographic characteristics [3]. Differences in cultural acceptance, population density, and accessibility to campaign materials and testing and care services could markedly affect intervention outcomes. Future work should focus on tailoring interventions to accommodate regional variations in HIV dynamics and barriers to screening and early diagnosis, ensuring more universally effective HIV prevention strategies.

Our study focused on analyzing HIV transmission dynamics, particularly with regard to new HIV infections originating within the Netherlands. Therefore, our analyses did not explore the implications of infections related to immigration or mobility of MSM. However, according to HIV surveillance data, the rate of new HIV infections originating within the Netherlands is decreasing. It is conceivable that within 10 years, the importation of new HIV infections may become a driving force in HIV transmission dynamics. This trend is already gaining attention in Denmark as demonstrated by a study by Palk et al. [37]. Consequently, the intervention aiming to increase the diagnosis of individuals with early infection may cease to be effective. Several reasons could contribute to this. For example, the engagement strategy may no longer be suitable for the target audience, considering both the delivery of information and the intended receiver’s ability to act on it. The latter could be influenced by factors such as perceived stigma, trust in, and access to healthcare [38]. This situation calls for the development of new, more tailored interventions aimed at addressing these barriers.

## Conclusion

Our study demonstrates that accelerating early-stage HIV diagnosis can significantly lower new infections among MSM, particularly in settings like the modern-day Netherlands, where UNAIDS’ 95-95-95 targets are met. Key to translating these findings into practice is tailoring strategies to the unique needs and conditions of the local population. Our modeling, which showed the impact of increasing diagnosis rates to desired levels, assumed high retention in care and optimal ART adherence. To achieve the impact that our study projected, essential factors including engaging MSM effectively, ensuring access to testing and immediate ART initiation, and supporting sustained ART adherence need to be in place. By conducting thorough needs assessments and engaging with stakeholders, we can devise targeted early diagnosis strategies, moving closer to eliminating HIV transmission.

## Supporting information

**S1 Appendix. Model structure and parametrization** Full description of the model structure and parametrization.

**S2 Appendix. Additional analyses** Additional analyses supplementing results presented in the main text and sensitivity analyses of the findings to perturbation of parameters and condition of immediate initiation of ART.

## Supporting information

Model structure and parametrization

Additional analyses

## Data Availability

All data produced in the present study are available upon reasonable request to the authors

## Acknowledgments

The authors gratefully acknowledge funding by the Aidsfonds Netherlands, grant number P-53902. We thank Peter Reiss, Wim Zuilhof, Marleen Werkman, Martin Bootsma, Michiel van Boven, Kim Romijnders, Maarten Reitsema, Maria Xiridou, Don Klinkenberg, and the Dutch HIV Association for useful discussions.

