## Supplementary material for "Impact of increased diagnosis for early HIV infection and immediate antiretroviral treatment initiation on HIV transmission among men who have sex with men in the Netherlands": Model structure and parametrization

### 1 Model and parametrization

In this section, we present a comprehensive description of the model, outline the procedures used to estimate parameter values, and provide the parameters themselves. For the sake of completeness in the presentation and ease of reading, we include some of the information given in the main text.

We consider the population of men who have sex with men (MSM) in the Netherlands. Although the estimated size of the MSM population in the Netherlands ranges from 200,000 to 300,000 individuals [1], simulations with this full population size would not be computationally tractable. Therefore, we use a population size of 25,000 individuals, which is both computationally tractable and sufficiently large to yield representative results that can be extrapolated to the national level.

In this study, we assess the impact of the intervention over 10 years. Within this period, three processes influence the epidemiological state of the population: demographic processes, sexual network dynamics, and HIV transmission dynamics. Demographic processes encompass the entrance and exit of individuals to and from the population, as well as ageing. Sexual network dynamics involve the formation and dissolution of sexual partnerships that facilitate contacts, leading to HIV acquisition among individuals exposed to the virus. Finally, HIV transmission dynamics encompass the natural progression of HIV in an exposed individual, in conjunction with care and prevention interventions, namely, diagnosis, the subsequent cascade of care, and a pre-exposure prophylaxis (PrEP) program.

The model is agent-based and, therefore, stochastic. Thus, to capture the full range of possible dynamics for a given scenario, an ensemble of trajectories from simulation runs is collected and subsequently analyzed. Each scenario corresponds to a fixed parameter set, and stochasticity arises from population dynamics. For each fixed parameter set, we create 400 simulation runs, each with a distinct random seed used by the random generator. To assess the impact of parameter perturbation on the model's projections, we conducted sensitivity analyses, generating 400 trajectories per parameter perturbation (see Section ??).

### 1.1 Simulation details

The simulation begins with a population of 25,000 individuals, progressing with constant one-day steps. At each step, demographic, sexual network, and HIV transmission events can occur based on the population-state-dependent probability. As events occur, the state of affected individuals is updated accordingly. The simulation run comprises three distinct stages: (a) An initial two-year burn-in stage, during which sexual network dynamics settle to a pseudo-equilibrium state without activating HIV processes; (b) The subsequent 6.5 years dedicated to establishing the epidemiological state of the population, where the infection profile of the population is initialized and evolves according to pre-defined rules. This stage of the simulation is used to calibrate the model by matching the evolution of HIV dynamics in the population of MSM during the 2017-2022 period; (c) The final 10-year stage during which intervention scenarios are modeled. The simulation timeline commences on July 1, 2014.

### 1.2 Demography

The demographic processes encompass the entrance of individuals into the population, the exit of individuals due to mortality unrelated to HIV dynamics (background mortality), and ageing.

In the population of MSM, HIV transmission predominantly occurs through condomless anal intercourse [2]. Therefore, we consider a population of sexually active men aged 15 to 74 years. Additionally, we classify the population by age into groups corresponding to 10-year age bands: 15-24, 25-34, 35-44, 45-54, 55-64, and 65-74 years.

At the start of the simulation, we initialize the population with characteristics reproducing trends observed in the data. In the context of demography, the age distribution of the model population mirrors the age distribution for the male population in the Netherlands in 2014 [3]. We use the discrete distribution obtained from data analysis to classify individuals into age groups. Subsequently, we assign the exact age of each individual by uniformly sampling the width of the age group to which they belong. Observe that age characteristic affects intrinsic properties of individuals such as the propensity to acquire non-steady sexual partners and background mortality rate.

All new individuals enter the population at the age of 15 years. While the population that we consider includes individuals who have immigrated to the Netherlands, we do not model the effects of immigration on the makeup of the population during the simulation.

Individuals leave the population once they reach the age of 75 years, simulating cessation of sexual activity. In addition to leaving the population after cessation of sexual activity, individuals can leave the population at any other moment as a result of background age-dependent mortality [4]. We model mortality within each age band using a geometric distribution process, using WHO life tables for the Netherlands [4] to calculate the mortality rates for each age group. In our model, individuals who leave the population are not being immediately replaced.

As the simulation moves forward in time, individuals age, such that everyone's age increases by 1 year every time the time counter passes a multiple of 365 days. Once an individual ages from one age band to the other, their mortality rate changes respectively. To model the appearance (or "birth") of new individuals in the population we assume that the birth of new individuals is a Poisson process with a mean of $\lambda$ individuals entering the population per year. This parameter is calculated using the expression for the population size at an equilibrium which is derived from the analogous deterministic model. This ensures a quasi-stable population size over the simulation run.

Parameters pertinent to demographic dynamics are summarized in Supplementary Table 1.

| Description (unit) | Value | Source |
|---|---|---|
| Time spent in each age band (years) | 10 | Follows from the definition of age bands |
| Background death rate in age band (year$^{-1}$) | | WHO life tables for the Netherlands [4] |
| 15-24 | $0.3 \times 10^{-3}$ | |
| 25-34 | $0.5 \times 10^{-3}$ | |
| 35-44 | $0.9 \times 10^{-3}$ | |
| 45-54 | $2.4 \times 10^{-3}$ | |
| 55-64 | $7.0 \times 10^{-3}$ | |
| 65-74 | $20.5 \times 10^{-3}$ | |
| Mean rate of entrance of new individuals (individuals year$^{-1}$) | 447 | Derived from the deterministic analog model using 25,000 individuals as an equilibrium population size |

Supplementary Table 1: **Demographic processes: summary of model parameters.**

#### 1.3 Sexual network dynamics

To accurately model the transmission dynamics of HIV in the population of MSM, the model captures the dynamic nature of the sexual network and its dependence on the demographic makeup. The model distinguishes between two types of partnerships: steady and non-steady. We use the term “steady partnership” in the meaning defined by EMIS-2010 [5] and EMIS-2017 [6] to mean a long-term partnership such that individuals who participate in such relationships do not consider themselves single. The term “non-steady partnership” refers to all other pairs who have sexual intercourse. These include one-time sex acts (with known and unknown partners), one-night stands (with known and unknown partners), relationships with more than one meeting but without an agreed-upon arrangement, and sex-buddy arrangements. The key difference between the two types of pairs is 1. duration of the relationship; 2. frequency of AI; 3. proportion of AI which are condomless; 4. concurrency.

For both types of partnerships, partner selection is influenced by age and HIV status, i.e., we account for age assortativity and serosorting.

Supplementary Table 2 lists sexual network dynamics parameters and their values.

##### 1.3.1 Steady partnerships

Analysis of EMIS-2017 data has shown that a large proportion of men who have sex with men indicated either that they had no steady male partner (approximately 55%) or were in a steady partnership with one man only (42%), only a small proportion indicated (3%) that at the time of the survey they were in a steady partnership with more than one man. Consequently, we model that, at any moment in time, an individual can either be single or in a steady partnership with one man. The simulation algorithm ensures that the proportion of individuals with a steady partner fluctuates around 45%.

At each point in time, in our model, a number of individuals can form new partnerships and a number of existing partnerships can dissolve. The number of new pairs formed per unit time depends on the number of available individuals (i.e. individuals without a steady partner) and the rate of steady partnership formation. The number of dissolved partnerships depends on the average duration of a steady partnership. We assume that each individual when single has the same rate of entering a steady partnership and that the duration of each partnership is on average the same for each pair, i.e., the duration of a partnership has a geometric distribution.

At the start of the simulation, a number of steady partnerships are formed, such that the quasi-equilibrium of 44% of individuals with a steady partner is maintained. This is achieved by fixing the partnership formation rate via calibration of the system's dynamics. For each pair, the first partner is selected randomly among individuals without a steady partner, then their age band and HIV status is accessed. We look up the age preference by age for this individual (Supplementary Table 3) and determine the age of the prospective partner by sampling a random process with a discrete number of outcomes (age bands of the partner). Similarly, the HIV status of the prospective partner is determined by sampling respective two-entry preference distribution (Supplementary Table 5). Once the age and the HIV status are determined, a second partner is sampled from a sub-population of individuals with selected age and HIV status and no steady partner. If two selected individuals are not currently involved in a non-steady partnership, a new pair is formed.

As the simulation progresses, new partnerships are formed such that the number of individuals with a steady partner fluctuates at the pre-determined level. When a partnership is created its duration is sampled. Throughout the simulation progress, the end dates of partnerships are checked whether the end date corresponds to the in-simulation date. For partnerships where this is true, the partnership is dissolved and individuals rejoin the pool of those without a steady partner.

This modeling paradigm is based on the pair-formation model introduced by Dietz and Hadeler [7] and subsequently extended [8] and used to model STI transmission and assess the impact of interventions on their dynamics by Kretzszhmar, Heijne and others [8, 9, 10]. Analysis of the deterministic analog shows that the number of individuals with a steady partner is expected to fluctuate around a steady state value, determined by the formation rate of the partnership and its average duration.

#### 1.3.2 Non-steady partnerships

In addition to steady partnerships, individuals can form non-steady partnerships. Unlike steady partnerships, individuals who have steady and non-steady partners can form new non-steady partnerships. The data indicates that the propensity to form such partnerships is heterogeneous with some individuals reporting very few (0-2) over 6 months, while others reporting much higher numbers ($> 6$) over the same period (Figure 1).

Available data from ACS indicates that most non-steady partnerships are brief, lasting just a few days. Therefore, the approach we took with modeling steady partnership formation is not a suitable fit for non-steady partnership formation, as due to their brevity, the non-steady partnerships for most individuals rarely overlap or have an effect on the acquisition rate of such partners. Instead, it is more organic to consider the possibility that each individual has a characteristic propensity for forming non-steady partnerships. Individuals with higher propensity have a higher chance of being recruited to form such partnerships than individuals with lower. Analysis of the ACS data yielded that the acquisition rate of non-steady partners depends on the age of the individual and whether they have a steady partner (Supplementary Figure 1).

Similar to the steady partnerships modeling paradigm, at each point in time individuals can form new non-steady partnerships and existing non-steady partnerships can dissolve.

The number of new pairs formed at each point in time depends on the number of individuals in the population and the formation rate of non-steady partnerships. If the simulation determines that a pair is to be formed, it recruits two potential partners from the population such that individuals with a higher propensity to form this type of partnership are picked with higher probability.

More precisely, when the simulation determines that a pair is to be formed, it selects the first partner, such that individuals with a higher propensity to form non-steady partnerships are chosen with higher probability. Once the first partner is chosen, their age and HIV status are evaluated. Based on these criteria, the distributions describing the tendency to select a partner with respect to age and HIV status are retrieved (Tables 4 and 5). Both of these discrete distributions are sampled, and the age and the HIV status of the second partner is determined. From the pool of individuals matching these characteristics, a second partner is chosen such that individuals with a higher propensity to form a non-steady partnership are more likely to be selected. If the two individuals are currently not engaged in a steady or non-steady partnership with each other a pair is formed. When a non-steady pair is formed the time of its dissolution is sampled from an exponential distribution. As the simulation progresses, the end date of each non-steady partnership is checked against the in-simulation date.

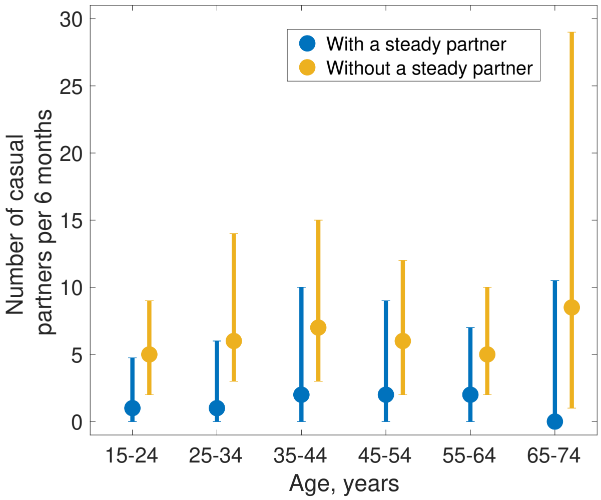


Supplementary Figure 1: **Distribution of acquisition rate of non-steady partners by age, with and without a steady partner.** Analysis of the rates of acquisition of non-steady partners per 6-month period using data Amsterdam Cohort Studies (ACS) [11]. Large dots denote medians of distribution with whiskers denoting quartiles.

To estimate rates of formation of steady and non-steady partnerships, we analyzed the data from the MSM Network Study (NS) in Amsterdam [12, 13] and estimated that a mean duration of a non-steady partnership is 2 days. From the analysis of the cross-sectional European MSM Internet Survey (EMIS) 2017 [14] data, we obtained an estimate of the proportion of the population having a steady partner, and the median number of steady partners within the last 12 months. From the analysis of the Amsterdam Cohort Study, we obtained the median number of non-steady partners within the last 6 months. The calibration of the model against these target characteristics used Nelder-Mead function optimization method, using of **fminsearch** function in MATLAB [15], to estimate the rates of formation of steady and non-steady partnership, as well as the expected duration of a steady partnership. The optimization method used parameter values derived from equilibrium values in the analog deterministic pair-formation model as a seed state. We modeled the time before acquiring a partner and the duration of partnerships as being exponentially distributed.

| Description (unit) | Value | Source |
|---|---|---|
| Mean duration of a steady partnership (year) | 1.64 | MSM Network Study in Amsterdam [12, 13] |
| Formation rate of steady partnerships ($year^{-1}$) | 0.57 | EMIS-2017 [6] |
| Mean duration of a non-steady partnership (day) | 2.0 | MSM Network Study in Amsterdam [12, 13] |
| Formation rate of non-steady partnerships ($day^{-1}$) | 0.02 | ACS [11] |

Supplementary Table 2: **Summary of model parameters and transition rates: sexual network dynamics.**

##### 1.3.3 Age assortativity and serosorting

To obtain age-dependent and sero-dependent mixing patterns in the population, we analyzed data collected in the MSM Network study [12, 13].

To calculate age-mixing matrices for steady and non-steady partnerships we used the method described in [16]. In short, this method allows the estimation of contact frequency between different age groups by employing the maximum likelihood method to estimate the mean number of contacts between individuals in different age bands, while correcting for reciprocity of contacts by modifying the log-likelihood function using weights of relative size of each age band with respect to the total population. Once the rates were calculated, we normalized the contact rates for each age band, to obtain age preference distributions for each age band (Tables 3 and 4).

| | Age of the individual | | | | | |
|---|---|---|---|---|---|---|
| **Age of partner** | 15–24 | 25–34 | 35–44 | 45–54 | 55–64 | 65–74 |
| 15–24 | 0.3724 | 0.1592 | 0.0616 | 0.0392 | 0.0395 | 0.0000 |
| 25–34 | 0.4563 | 0.4866 | 0.3197 | 0.2392 | 0.2057 | 0.1470 |
| 35–44 | 0.1539 | 0.2811 | 0.4555 | 0.4478 | 0.3935 | 0.4526 |
| 45–54 | 0.0174 | 0.0589 | 0.1538 | 0.2465 | 0.2813 | 0.4004 |
| 55–64 | 0.0000 | 0.0105 | 0.0094 | 0.0245 | 0.0700 | 0.0000 |
| 65–74 | 0.0000 | 0.0037 | 0.0000 | 0.0028 | 0.0100 | 0.0000 |

Supplementary Table 3: **Age-dependent mixing preference for individuals in steady partnerships.**

| Age of partner | Age of the individual | | | | | |
|---|---|---|---|---|---|---|
| | 15–24 | 25–34 | 35–44 | 45–54 | 55–64 | 65–74 |
| 15–24 | 0.4002 | 0.1633 | 0.0662 | 0.0377 | 0.0497 | 0.1036 |
| 25–34 | 0.4184 | 0.5101 | 0.3670 | 0.2409 | 0.1972 | 0.1363 |
| 35–44 | 0.1306 | 0.2753 | 0.4489 | 0.4705 | 0.4444 | 0.3216 |
| 45–54 | 0.0421 | 0.0428 | 0.1082 | 0.2253 | 0.2499 | 0.3343 |
| 55–64 | 0.0086 | 0.0066 | 0.0097 | 0.0242 | 0.0588 | 0.1042 |
| 65–74 | 0.0000 | 0.0019 | 0.0000 | 0.0014 | 0.0000 | 0.0000 |

Supplementary Table 4: **Age-dependent mixing preference for individuals in non-steady partnerships.** Each column is normalized to add up to 1.

To identify tendencies of MSM to serosort, we analyzed the MSM Network Study data. In this study, the respondents reported the knowledge or perception of their own HIV status, as well as the knowledge or perception of the HIV status of their partner. Based on the data, and adjusting for the prevalence of individuals with diagnosed HIV infection in the population of MSM, we have calculated matrices describing the probability distribution of sexual mixing for steady and non-steady partnerships (Supplementary Table 5). In effect, the serosorting delineates the population into two subgroups: those who have received an HIV diagnosis and those who have not.

| | Steady partnerships | | Non-steady partnerships | |
|---|---|---|---|---|
| | **No HIV diagnosis** | **HIV diagnosed** | **No HIV diagnosis** | **HIV diagnosed** |
| **No HIV diagnosis** | 0.981 | 0.254 | 0.982 | 0.236 |
| **HIV diagnosed** | 0.019 | 0.746 | 0.178 | 0.764 |

Supplementary Table 5: **Serosorting patterns distributed by the type of the partnership.** Each column is normalized to add up to 1.

### 1.4 HIV dynamics

In this section, we describe in detail the modeling process of the natural history of HIV transmission in MSM coupled with care and prevention interventions which were ongoing in the Netherlands in 2023.

#### 1.4.1 Natural progression of infection

To model the natural history of HIV without medical interventions, such as testing, and enrolling in antiretroviral therapy (ART) and pre-exposure prophylaxis (PrEP) programs, we use a standard framework. In this model, susceptible individuals are exposed to HIV through condomless anal intercourse

(AI) with an infectious partner, potentially leading to HIV acquisition. Individuals who newly acquired HIV infection enter a brief early HIV infection stage, followed by a prolonged chronic HIV infection stage, which culminates in the AIDS stage. We also divide the AIDS stage into two sub-stages: the early AIDS stage and the late AIDS stage. In all stages following the transmission event, individuals with HIV may transmit. Conversely, susceptible individuals get infected at a rate that is proportional to the number of infectious individuals within the local sexual network of the susceptible individual.

The early stage has a short duration (in the model an average of 89 days [17]), which is characterized by elevated viral load reflected in the temporary increased probability of transmitting HIV. Following the early infection stage, individuals enter the chronic infection stage where their health profile, as reflected by viral load, remains relatively constant, and clinical manifestations of the infection are little to none. In the model, this stage on average lasts for 8.31 years [9], after which period individuals enter the early AIDS stage, with opportunistic infections and AIDS-related oncology diseases. After an average of 1.18 years [9] individuals enter the final stage of infection, the late AIDS stage, characterized by a rapid rise in the viral load and onset of multi-system diseases. Both early and late AIDS stages are characterized by additional mortality rates, with life expectancy after entering the AIDS stage equal to 3 years [9]. To ensure that the duration of the early stage of HIV infection does not end exceedingly quickly, in the model we split this period into three substages, corresponding to Fiebig stage 1, Fiebig stages 2 and 3, and Fiebig stages 4 and 5 [17] with mean duration equal to 5, 9, and 75 days, respectively. The time spent in each substage is modeled using an exponentially distributed random process. We model the duration of AIDS stages using exponentially distributed random processes, while the duration of the chronic stages is modeled with Erlang distribution with shape parameter 80, to restrict the variance.

As individuals progress through the stages of HIV infection, their viral load changes. Thus, the early HIV infection stage is characterized by a sharp increase and subsequent decrease of the viral load, corresponding to a significantly higher potential for HIV transmission per condomless anal intercourse event compared to the chronic HIV infection stage. Similarly, individuals in the AIDS stage of infection have a greater infection transmission potential than those in the chronic stage. To reflect this dynamic, we assigned varying probabilities of transmission to individuals in different HIV infection stages. Based on the estimates from the literature [9], we set the chronic HIV infection stage as a baseline, the potential to transmit in the early stage of HIV infection is 26 times higher, while in the early AIDS stage, the potential is increased by a factor of 6. While individuals in the late AIDS stage have a still higher potential to transmit, the significant AIDS-associated morbidity leads us to adopt the abstraction that such individuals do not engage in sexual activity. The probability of transmission in the chronic stage of HIV infection was estimated by calibrating the model to the annual numbers of new HIV infections and HIV diagnoses.

The probability of transmission per day is a composite of the rate with which anal intercourse takes place, the probability of condoms being used, and the probability of HIV transmission per condomless anal intercourse. To estimate the average number of contact rates in different partnerships, we analyzed the data collected in the MSM Sexual Network survey [12, 13]. A steady partnership has an average contact rate of 0.33 contacts per day. Non-steady partnerships are characterized by the mean AI rate of 0.11 contacts per day. Moreover, to ensure that each such partnership has at least one AI during its course, we modeled that each non-steady partnership starts with an AI.

To estimate the probability of condom use, we analyzed the proportion of steady and non-steady partners with whom respondents of EMIS-2017 data set [14] reported having condomless anal intercourse. Analysis of the data set yielded that condom use during anal intercourse differed across different types of partnerships, ages of individuals, and their HIV status (Supplementary Table 6). Since the number of respondents diagnosed with HIV and aged 15-24 and 65-74 was too low to be representative, we extrapolated the values in the adjacent age groups, 25-34 and 55-64, respectively. When simulating an anal intercourse event in a partnership, we reconcile condom use by taking the highest value between the two partners.

| Partnership Type | Age of the individual | | | | | |
|---|---|---|---|---|---|---|
| | 15–24 | 25–34 | 35–44 | 45–54 | 55–64 | 65–74 |
| **Steady (HIV-negative/not diagnosed)** | 0.29 | 0.22 | 0.28 | 0.28 | 0.28 | 0.27 |
| **Steady (HIV-positive/diagnosed)** | 0.1 | 0.1 | 0.14 | 0.17 | 0.16 | 0.16 |
| **Non-Steady (HIV-negative/not diagnosed)** | 0.63 | 0.65 | 0.58 | 0.54 | 0.55 | 0.48 |
| **Non-Steady (HIV-positive/diagnosed)** | 0.29 | 0.29 | 0.27 | 0.24 | 0.34 | 0.34 |

Supplementary Table 6: **Condom use probability in steady and non-steady partnerships.**

#### 1.4.2 Care and prevention programmes

**Diagnosis and cascade of care.** As described previously, individuals who have acquired HIV infection can be diagnosed and subsequently start taking treatment with the ultimate goal of achieving an undetectable viral load. The rate at which individuals with HIV infection get diagnosed depends on the HIV infection stage. In the simulation, we fixed the average diagnosis rate for individuals in the AIDS stage to be equal to a rate of 1/12 per year. This relatively high rate is meant to mimic the appearance of AIDS-related illnesses, which may prompt individuals in this stage of HIV infection to seek a diagnosis. We model the diagnosis process in each stage as exponentially distributed random processes.

To determine the diagnosis rates for various stages of HIV infection—including early HIV infection, chronic HIV infection in individuals who acquired the infection more than three months but less than six months ago, chronic HIV infection in those who acquired it more than six months ago but less than one year ago, and chronic HIV infection in individuals who acquired the infection more than 12 months ago—we aligned the model's outputs with data from Stichting HIV Monitoring (SHM). This organization tracks HIV trends in the Netherlands. We used the data detailing the estimated annual number of new HIV infections, the annual number of new HIV diagnoses, and the annual distribution of the stage of HIV infection at the time of diagnosis. Through the calibration process, we determined that the diagnosis rates for individuals with early HIV infection and those with chronic HIV infection acquired less than six months prior were nearly identical and higher compared to the diagnosis rates for individuals with chronic HIV infection acquired more than six months but less than twelve months ago. This, in turn, was higher than the diagnosis rate for individuals with chronic infection for more than twelve months.

Following diagnosis, individuals start ART treatment and subsequently can achieve viral suppression. Based on SHM data, we modeled the time between the diagnosis and ART initiation as an exponential process with a mean of 2 months. While the transition period from receiving an HIV diagnosis to starting treatment is longer than what is currently observed, it is shorter than estimates from 2010 (page 17, Chapter 1, [18]). This duration was chosen to align the model outputs with reported percentages of diagnosed individuals who have initiated treatment. The time from ART initiation to suppression was similarly modeled to be exponentially distributed with a mean of 2 months. The rationale for this value stems from the results by Dijkstra et al. [19] who have shown that individuals detected in the early stages of HIV infection and who immediately initiated diagnosis within 24 hours of diagnosis reached suppression with a median time of 55 days (IQR: 31–72). When an individual receives an HIV diagnosis, we model the contact rates in both types of partnerships as reduced by half. This contact reduction lasts until the individual achieves viral suppression.

Finally, a fraction of individuals can drop out of treatment or may experience treatment failure, i.e., the viral load is no longer suppressed. When dropping out of treatment voluntarily, we assume that individuals move to the stage corresponding to individuals with chronic HIV infection who were diagnosed. For simplicity, we assume that these individuals can be offered treatment again. When modeling the failure of viral suppression, we assume that individuals promptly get started on a different line of treatment and once again achieve viral suppression [18]. We determined the ART failure/dropout rate by calibrating the model outputs to the available data. The data involved the following three annual proportions: 1. diagnosed individuals out of the overall population living with HIV, 2. individuals who started ART relative to the overall population who received an HIV diagnosis, and 3. individuals who achieved viral suppression relative to individuals who started ART.

**PrEP.** In the Netherlands, a national PrEP program oriented towards people at risk of acquiring HIV, of whom MSM are a substantial portion, started in 2019. Before this time, PrEP uptake was low and those who used it either procured it from abroad or received it via the AMPrEP demonstration study which ran in Amsterdam, the Netherlands, from 2015 to 2020. In addition to the national PrEP program, PrEP may be obtained via general practitioners. As of the time of writing of this manuscript, the number of MSM currently enrolled in the PrEP program was capped at 9,000; thus, if we assume a population of 200,000 individuals, i.e., an estimated 4.5% used PrEP. We modeled PrEP uptake as starting with no users at the end of 2018, PrEP use starting in 2019, and gradually growing towards a level of 4.5% by January 2022, after which time the the percentage of PrEP users fluctuates around this value. While PrEP can be taken either as a daily preparation or timed around the anticipated time of sexual activity, for simplicity we modeled the use as continuous.

The general process of participation in the PrEP program is as follows: as the simulation progresses, at each step, a number of individuals who correspond to the criteria for PrEP program enrollment start using PrEP. Similarly, individuals who used PrEP can leave the program. Both enrollments to and leaving the program are modeled using exponential waiting times. Since the idea behind the PrEP program is to distribute it to individuals most at risk of HIV acquisition, we approximated the PrEP eligibility criteria using the number of non-steady partners within the last six months. Individuals who had 5 or more partners, were eligible to enroll. At the time when our study was performed, the PrEP program was active for approximately 3 years, and the average duration of being in the program would be hard to estimate without encountering censuring bias, therefore we assumed an average of 2 years and a possibility to enroll to the program several times. Individuals who use PrEP, provided they have a high adherence to the regimen, have a lower probability of acquiring HIV during a condomless anal intercourse act. Based on the available evidence, we set the relative reduction in the risk of acquisition to be 86% [20].

If an individual using PrEP has acquired HIV infection, then, adopting a modeling abstraction, this individual is considered to have started ART treatment upon the infection event. This abstraction captures the medical standard where PrEP users undergo regular HIV and STI testing.

#### 1.5 Initialization and simulation run

At the start of the simulation, we initialize the age distribution of the population based on the general male population in the Netherlands in 2014 [3]. Subsequently, we initialize a number of steady partnerships to ensure the proportion of individuals in steady partnerships aligns with data in EMIS-2017 [6]. We then determine the likelihood of individuals forming non-steady partnerships, considering the individual's age and whether they are in a steady partnership. After completing this initialization, the model runs for 2 years, simulating only demographics and sexual network dynamics, to achieve a (pseudo)-equilibrium state in sexual network dynamics. At this point, the calibration process starts as we initialize the HIV epidemic state to reflect 2016 statistics, including the total number of individuals living with HIV, the percentage diagnosed, those who have initiated ART treatment, and those achieving viral suppression [21]. For each simulation trajectory, the total number of individuals living with HIV is uniformly sampled within the confidence interval provided by SHM for 2016. The specific proportions for diagnosis and cascade of care were set to fixed values equal to SHM's exact values for that year. The model then continues to simulate demographic, sexual network, and HIV transmission dynamics. By the end of what would be 2019, the simulation of the PrEP program is activated. Following the calibration process, which concludes at the end of 2022, the main simulation begins. Depending on the scenario, this phase may include increased diagnosing of individuals with early HIV infection or those who acquired the infection in the previous 6 months.

#### 1.6 Calibration

To calibrate the model, we identified a feasible parameter space and sampled it using Latin Hyper Cube Sampling [22] using 800 parameter sets. For each set, we created an ensemble of 40 stochastic trajectories, which were summarized using appropriate statistics. We assessed the goodness of fit using the following metric:

$$d\left(v^{\mathrm{t}}, v^{\mathrm{s}}\right) = \max_i \frac{\left|v_i^{\mathrm{t}} - v_i^{\mathrm{s}}\right|}{v_i^{\mathrm{t}}} \ , \tag{1}$$

where $v^{\mathrm{t}}$ denotes a target statistic and $v^{\mathrm{s}}$ denotes the respective simulation output. Observe that this metric is non-dimensionalized, facilitating the goodness of fit assessment across statistics with different dimensions. We selected the parameter set which minimized the sum of distances from the target statistics. The comparison between the data and the model outputs is shown in Figure 1 in the main text.
