## Additional analyses for "Impact of increased diagnosis for early HIV infection and immediate antiretroviral treatment initiation on HIV transmission among men who have sex with men in the Netherlands"

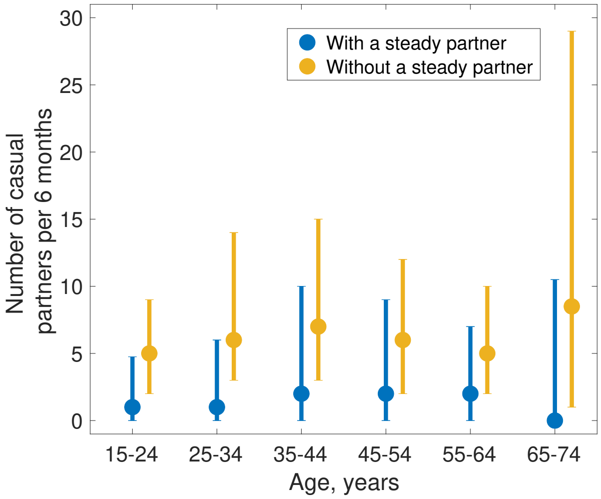


Supplementary Figure 1: **Distribution of acquisition rate of non-steady partners by age, with and without a steady partner.** Analysis of the rates of acquisition of non-steady partners per 6-month period using data Amsterdam Cohort Studies (ACS) [11]. Large dots denote medians of distribution with whiskers denoting quartiles.

| | **Steady partnerships** | | **Non-steady partnerships** | |
|---|---|---|---|---|
| | **No HIV diagnosis** | **HIV diagnosed** | **No HIV diagnosis** | **HIV diagnosed** |
| **No HIV diagnosis** | 0.981 | 0.254 | 0.820 | 0.236 |
| **HIV diagnosed** | 0.019 | 0.746 | 0.180 | 0.764 |

Supplementary Table 5: **Serosorting patterns distributed by the type of the partnership.** Each column is normalized to add up to 1.
